# User feedback on the NHS Test & Trace Service during COVID-19: the use of machine learning to analyse free-text data from 37,914 UK adults

**DOI:** 10.1101/2022.11.24.22282691

**Authors:** P Bondaronek, T Papakonstantinou, C Stefanidou, T Chadborn

**Affiliations:** Office for Health Improvement & Disparities, Department of Health and Social Care, London, United Kingdom, SW1H 0EU; Institute of Health Informatics, University College London, London, United Kingdom, NW1 2DA

## Abstract

**Objectives:** The UK government’s approach to the pandemic relies on a test, trace and isolate strategy, mainly implemented via the digital NHS Test & Trace Service. Feedback on user experience is central to the successful development of public-facing services. As the situation dynamically changes and data accumulate, interpretation of feedback by humans becomes time-consuming and unreliable. The specific objectives were to 1) evaluate a human-in-the-loop machine learning technique based on structural topic modelling in terms of its serviceability in the analysis of vast volumes of free-text data, 2) generate actionable themes that can be used to increase user satisfaction of the Service.

**Methods:** We evaluated an unsupervised Topic Modelling approach, testing models with 5-40 topics and differing covariates. Two human coders conducted thematic analysis to interpret the topics. We identified a Structural Topic Model with 25 topics and metadata as covariates as the most appropriate for acquiring insights.

**Results:** Results from analysis of feedback by 37,914 users from May 2020 to March 2021 highlighted issues with the Service falling within three major themes: multiple contacts and incompatible contact method and incompatible contact method, confusion around isolation dates and tracing delays, complex and rigid system.

**Conclusions:** Structural Topic Modelling coupled with thematic analysis was found to be an effective technique to rapidly acquire user insights. Topic modelling can be a quick and cost-effective method to provide high quality, actionable insights from free-text feedback to optimize public health services.

## Introduction

NHS Test & Trace is a crucial component of the UK’s government’s COVID-19 recovery strategy [1]. The Test & Trace Service traces the contacts of COVID-19 positive cases, and provides guidance on next steps for both cases and contacts. The Service comprises of a web-based, phone contacts, emails and text messaging. However, adherence to all parts of the process have been poor [2]. Improving how the NHS Test & Trace Service works for users is a necessary part of solving this problem.

Feedback on user experience is central to the successful development of public-facing services, no less so when systems have been developed rapidly in a pandemic [3]. If individuals have a negative experience with the Service, they may not complete data input, be less likely to use it in future, and more likely to report negative experiences to others. In addition, not believing that contact tracing systems are effective predicts not engaging with them [4].

Users of the Test & Trace Service are asked to rate their experience and leave feedback on how the system can be improved. Large volume of free-text data has been gathered with >1,800,000 words were produced up to March 2021 just by those who said they were dissatisfied/very dissatisfied. Human coders have only been able to examine a portion of the data (0.04% of the data collected from May to October 2020).

Such data requires an automated approach that is enabled by Artificial Intelligence (AI) and Machine Learning (ML) approaches. Topic modelling is an unsupervised Natural Language Processing (NLP) approach that has gained popularity in the analysis of free text data. Topic models are generative models of words counts, that are able to automatically infer latent topics from text. They are particularly useful as tools to analyse large volumes of open-ended free-text responses in an exploratory bottom-up manner. Our approach is based on an application of the Structural Topic Model [5] in particular. The STM is a general framework for topic modelling that enables the researchers to include document-level metadata as covariates in a topic model and estimate their relationships to specific topics.

During the course of the pandemic topic modelling has been used in to help provide insights on public’s behaviours and attitudes on COVID-19 and the various measures put in place. Some examples of relevant applications are: understanding COVID-19 concerns and attitudes as expressed in social media and online forums [6-8]; categorising disinformation and mistrust [9]; surveillance of non-pharmacological interventions across the globe [10]; monitoring the content of news articles about COVID-19) [11]; and analysis of attitudes on mask wearing [12]. In all instances topic modelling has proved to be an excellent tool to acquire rapid, high-level insights on the various issues explored.

A study by Guetterman and colleagues [13] explored the use of NLP as a tool to augment and validate qualitative analysis. They found an NLP clustering technique base on the Wu-Palmer similarity measure [14] to be suitable both for providing a foundation for speeding up human qualitative analysis, and for validating the human analysis. Similarly, another recent study [15] found topic modelling and graphic clustering of free-text data and to be an effective assistive technique to human qualitative coding.

There is a need to develop a method to accelerate the understanding of the complexity of what influences human behaviours. Indeed, human behaviour is central to transmission of SARS-Cov-2, and changing behaviour is crucial to preventing transmission. Automated approaches to data analysis of instructed data is need to respond quickly to the changing situation. Moreover, other pandemic-related digital services face similar problems of rapid rollout and need rapid analysis of user feedback.

## Aims and objectives

The aim of this study was to evaluate the use of ML as a tool to help understand the issues with the NHS Test & Trace Service as expressed in the free-text user feedback responses. The specific objectives were to 1) evaluate the application of an unsupervised machine learning technique, 2) create actionable themes that can be used to increase user satisfaction of the Test & Trace Service.

## Methods

### Data

User feedback has been collected since the start of the service, on 28 May 2020. We are using data from that date until 7 March 2021. Because of time constraints and the pragmatic nature of this study as a methodological proof of concept, we did not include more recent data in the reported analysis. This feedback is collected after users complete the form, where they are asked to provide a satisfaction rating for the service they received, with options ranging from “Very satisfied” to “Very dissatisfied”. They are also asked “How could we improve this service?”, to which they can respond in free text. The responses to this question provide a rich dataset of recommendations that can be used to improve the Service to better cater to its users’ needs and respond to a dynamic situation.

On 7 March 2021 a total of 153,128 responses were collected. In this study we are only analysing the free-text responses from users who stated they are dissatisfied with the Service (64,492 responses). We made the decision to only analyse this subset of responses in order to more easily and quickly identify those aspects of the service users were most often dissatisfied with and minimise the amount of noise in the dataset.

We also have more structured data, such as whether the user is a case (person testing positive for SARS-CoV-2)/contact (person named by a case as potentially exposed), the region where they are based, tier and demographic information. The tier variable refers to the user journey through the contact tracing system (automated/call handler-case/contact).

### Text analysis procedure

#### Preprocessing

The original dataset includes data collected since the Service was first implemented in May 2020, up until March 2021, from users in England. We preprocessed the data using R (version 3.5.2), and cleaned the free text responses using base.R functions, the quanteda [[16] and stm [5] packages. We deleted observations with missing values and duplicate data and prepared the covariates for analysis. After preprocessing, the remaining data available for analysis included records of 37,914 users. These data were converted into token units using the quanteda package after punctuation and numbers were removed. Data preprocessing was completed by deleting stop words and stemming the tokens.

#### Structural topic model

Prior to running the models we run diagnostics to identify the optimal number of topics, according to both the relevant metrics and this analysis’ aims, focusing on the trade-off between semantic coherence and exclusivity [17]. We evaluated an unsupervised Topic Modelling approach, testing models with 5-40 topics and differing covariates in terms of coherence, residuals and interpretability by human coders (see https://osf.io/cvzd3/). We identified a Structural Topic Model with 25 topics and metadata as covariates as the most appropriate for acquiring insights.

#### Output

The output analysed consisted of lists of 8 stemmed keywords with different weightings which is what the model interprets as the ‘topic’ and 10 representative quotes for each topic. Different types of word weightings were generated with each topic where the following two types were analysed in subsequent qualitative analysis: 1) Highest Prob (words within each topic with the highest probability) and 2) FREX (words that are both frequent and exclusive, identifying words that distinguish topics). In addition to that a list of the most representative quotes for a given topic was generated by the algorithm. See Figure 1 for an example of the output.

**Figure 1.**
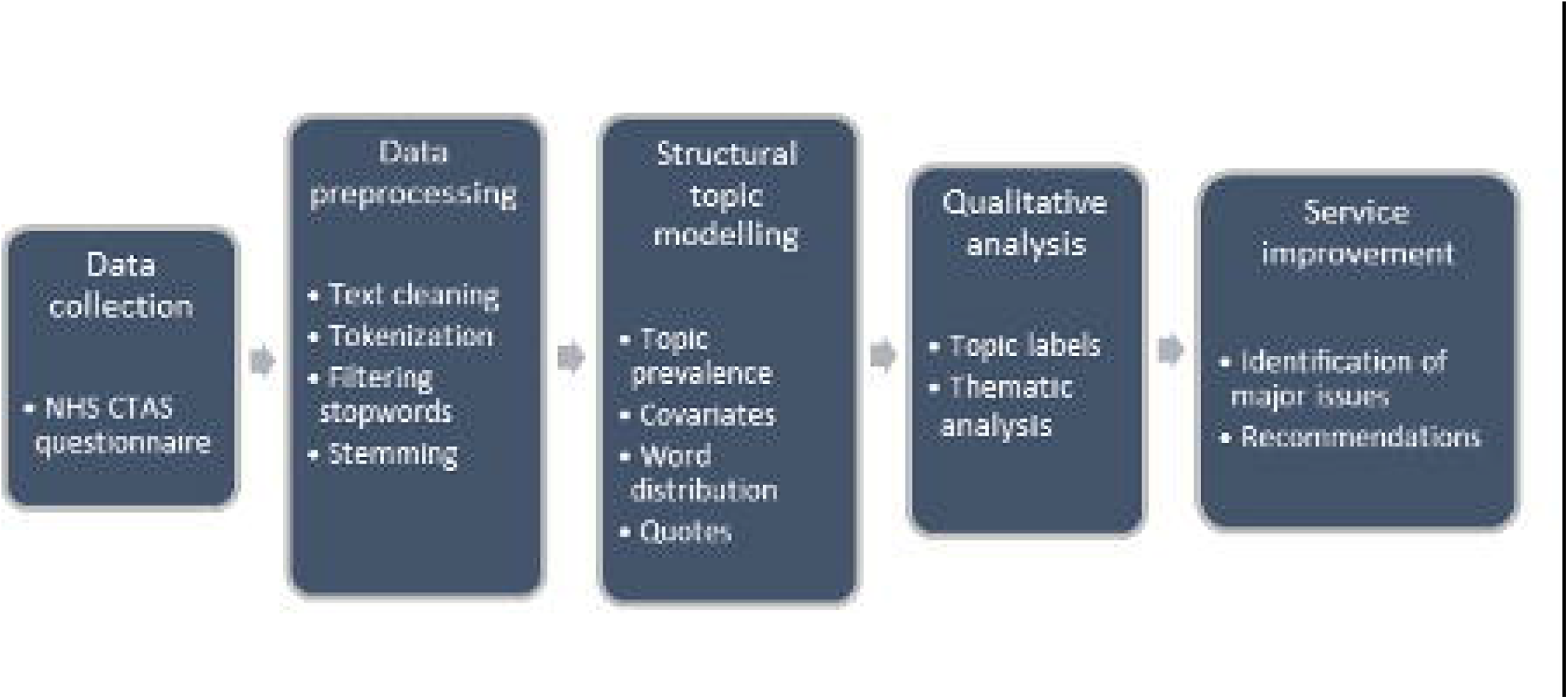
Example of the Structural Topic Model output

### Qualitative analysis process

The model’s output systematically was analysed systematically in two stages. In Stage 1, the descriptive titles for the topics were generated and agreed. In Stage 2, the six stages process of conducting a thematic analysis was adapted to guide to analyse the topics generated by the text analysis [18]. Two researchers (PB, TP) conducted the initial coding of the topics generated through test analysis. The process is summarised below.

First, the researchers familiarised themselves with the data through repeatedly reading the themes with associated quotes whilst searching for patterns. For this analysis, the emphasis was on identifying the issues that users of the Test & Trace Service were unsatisfied with. Secondly, the initial descriptions of the topics were generated. Third, the researchers identified the initial themes and discussed the similarities and discrepancies between them. Fourth, the researchers reviewed the themes whilst looking for any user feedback that might have been present in the topics identified but not represented in the major themes. This was a continuous process that was reviewed and re-defined (step five). Lastly, the report summarising the themes with illustrative quotes was generated (step six).

## Results

### Descriptive Statistics

Descriptive statistics for respondents are displayed in Table 1. The sample of dissatisfied users is majority female, with 57% falling within the 30-59 age group, evenly split between contacts and cases. Finally, the overwhelming majority completed the procedure entirely online.

**Table 1.**
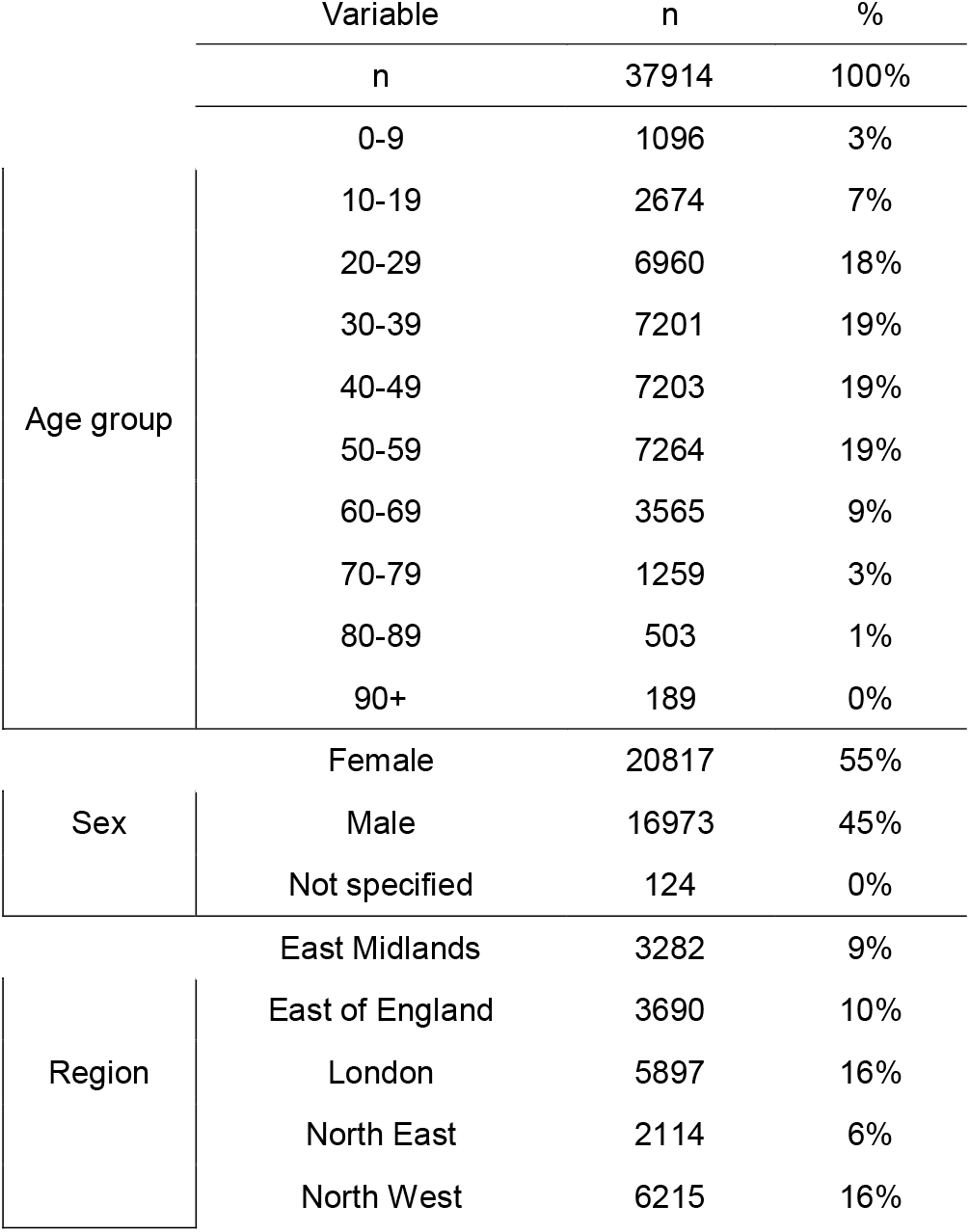
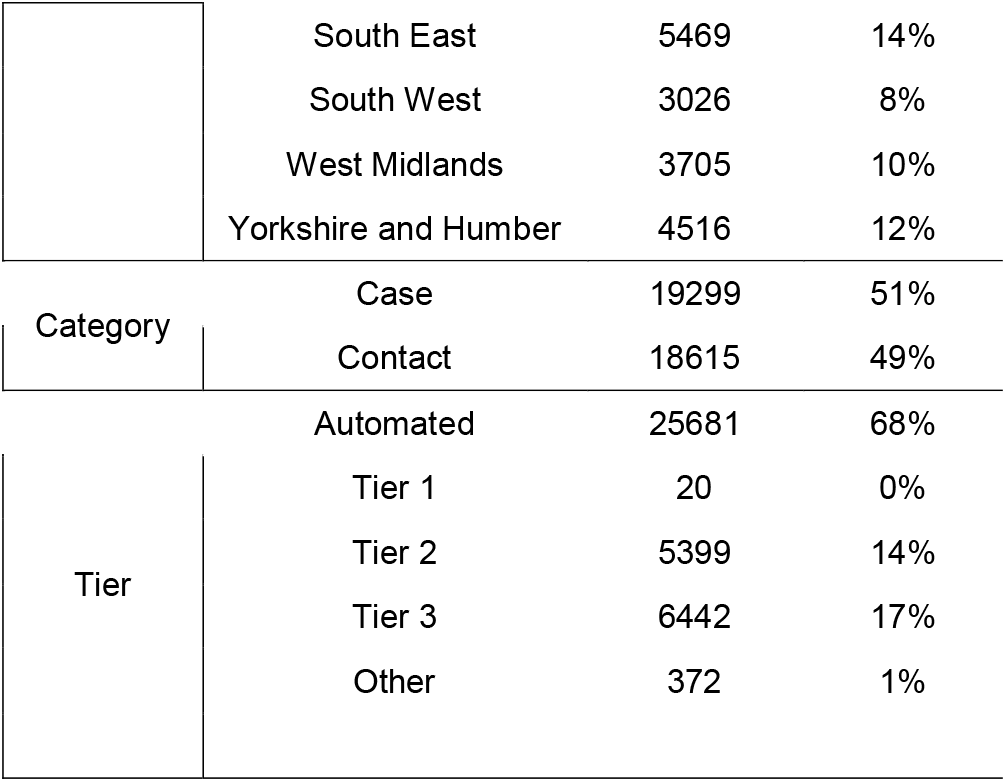
Sample description

|  |  | Variable | n | % |
| --- | --- | --- | --- | --- |
|  |  | n | 37914 | 100% |
| Age group |  | 0-9 | 1096 | 3% |
|  |  | 10-19 | 2674 | 7% |
|  |  | 20-29 | 6960 | 18% |
|  |  | 30-39 | 7201 | 19% |
|  |  | 40-49 | 7203 | 19% |
|  |  | 50-59 | 7264 | 19% |
|  |  | 60-69 | 3565 | 9% |
|  |  | 70-79 | 1259 | 3% |
|  |  | 80-89 | 503 | 1% |
|  |  | 90+ | 189 | 0% |
| Sex |  | Female | 20817 | 55% |
|  |  | Male | 16973 | 45% |
|  |  | Not specified | 124 | 0% |
| Region |  | East Midlands | 3282 | 9% |
|  |  | East of England | 3690 | 10% |
|  |  | London | 5897 | 16% |
|  |  | North East | 2114 | 6% |
|  |  | North West | 6215 | 16% |

|  |  |  |  |
| --- | --- | --- | --- |
|  | South East | 5469 | 14% |
|  | South West | 3026 | 8% |
|  | West Midlands | 3705 | 10% |
|  | Yorkshire and Humber | 4516 | 12% |
| Category | Case | 19299 | 51% |
|  | Contact | 18615 | 49% |
| Tier | Automated | 25681 | 68% |
|  | Tier 1 | 20 | 0% |
|  | Tier 2 | 5399 | 14% |
|  | Tier 3 | 6442 | 17% |
|  | Other | 372 | 1% |

### Inclusion of the topics in the qualitative analysis

Of 25 topics analysed qualitatively, 19 topics were included in the analysis as they provided substantial insights on various issues as expressed by the users’ feedback. The full output of the topic analysis is available in the online repository (https://osf.io/cvzd3/).

The rationale for exclusion of 6 topics from the analysis is provided below:

- Topic 22 related to the feedback on the testing service, specifically comments on inconclusive and inconsistent results, delay/not receiving tests, comments on perceived lack of the need to self-isolate due to negative results following positive one. Although this topic was important, it related to a separate issue than the Test & Trace Service per se
- One topic (topic 1) was deemed as incoherent due to the text analysis based on the Service name which was used repeatedly in the free text, i.e., “track” and “trace” and was excluded from further analysis
- Four topics included mixed issues that were represented in other themes (topics 11,5,3,6).

These remaining 19 topics were grouped into three major themes repressing the family of issues as expressed by the users of Contact Tracing (Figure 2):

**Figure 2.**
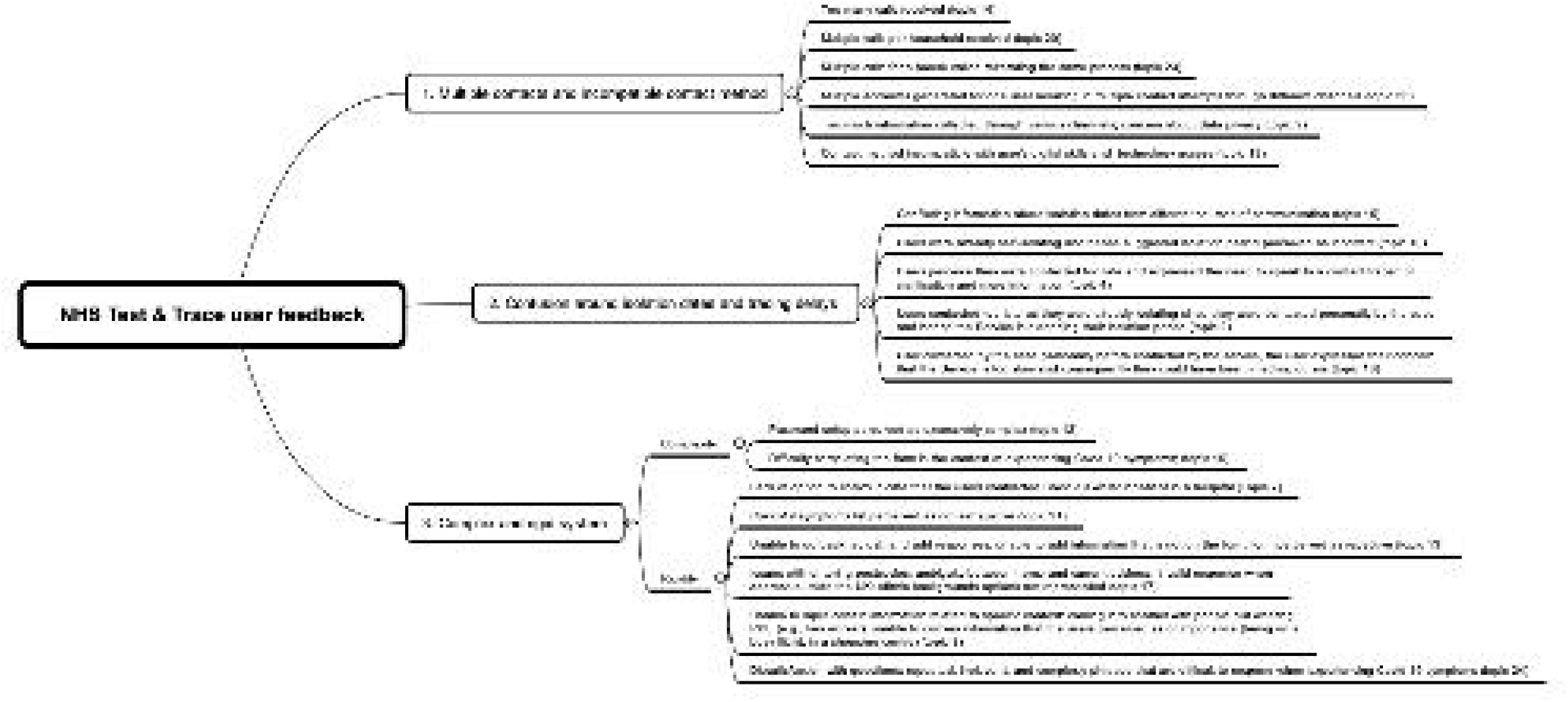
Visualisation of the qualitative analysis of the 19 topics

#### Major theme 1: Multiple contacts and incompatible contact method

The feedback expressed in this major theme related to the distress with receiving too many calls, especially in the context of experiencing Covid-19 symptoms. Users complained about multiple contacts per household reiterating the same process and expressed their views that the households should be ‘linked’ together. Users also perceived that too much information was collected, and there was some indication about data privacy concerns. In addition, participants expressed that multiple accounts were generated for one person, and hence they experienced multiple contact attempts through different channels, i.e., calls, emails and messages.

One topic related to the issue where the contact method did not match users’ digital skills and preferences. For example, users who did not own a smartphone were unable to access the links in the text message; those who did not have internet access; or were unable to fill in the online form due to lower digital skills.

#### Major theme 2: Confusion around isolation dates and tracing

The feedback expressed in this major theme related to receiving numerous and often conflicting information about isolation dates from different sources of communication. In many cases, users were already contacted by the case and started their isolation period. They expressed their concern about their perceived delay of the contact attempts by the Service because they “could have been carrying on as normal & spreading the disease”.

#### Major theme 3: Complex and rigid system

The feedback expressed in this major theme related to the complexity of the online form, especially around the difficulties with setting appropriate passwords and issues with completing the form in the context of experiencing Covid-19 symptoms. The form was seen as repetitive and rigid, with inability to go back and adjust responses. Some questions were seen as repetitive, irrelevant and phrased in a complex manner.

There were various issues with the logic flow of the form, and the inability to input information that was perceived as important. For example, the list of symptoms was perceived as not exhaustive, there were issues with entering postcodes, ambiguity between home and current address, and error messages when inputting addresses outside the UK, as well the lack of representativeness of certain ethnic backgrounds. The users perceived the options for information input as limited and, at the same time, some input requests were seen as repetitive.

## Discussion

### A statement of overall findings

This study provides some important insights into user experience of the Test & Trace Service as expressed through the free-text feedback survey based on over 37 thousand responses. The main issues related to multiple contacts attempts, delays in contact tracing, and technical issues with the form, specifically the complexity and rigidity of the survey. In line with previous research, technical issues were found to be a major cause of distress for users of the services, making their engagement with the Service challenging and, as a consequence, affecting their ability to adhere to guidance [19]. Incompatibility of contact method with level of digital skills was also shown to be a major issue expressed by users. Another important finding showed that users reported feeling overwhelmed and harassed by the number of contact attempts from contact tracers. This is in line with previous findings by Wright and colleagues [20].

Overall, Structural Topic Modelling was found to be an effective technique to rapidly acquire user insights. The 25 topics provided highly specific insights of issues that can be utilized towards improving the Test & Trace Service, with minimal human involvement and low maintenance requirements, making them ideally suited as tools for the evaluation of pandemic response services.

This study provides a proof of concept of a method to analyse large quantities of free text data in service improvement, combining an AI-enabled text analytics followed by a qualitative analysis conducted by a human. As such, this method has the potential to be to contribute to improvement of the Test & Trace Service, and other digital systems for public health safety or health improvement. This framework is represented in the diagram below (Figure 3).

**Figure 3.**
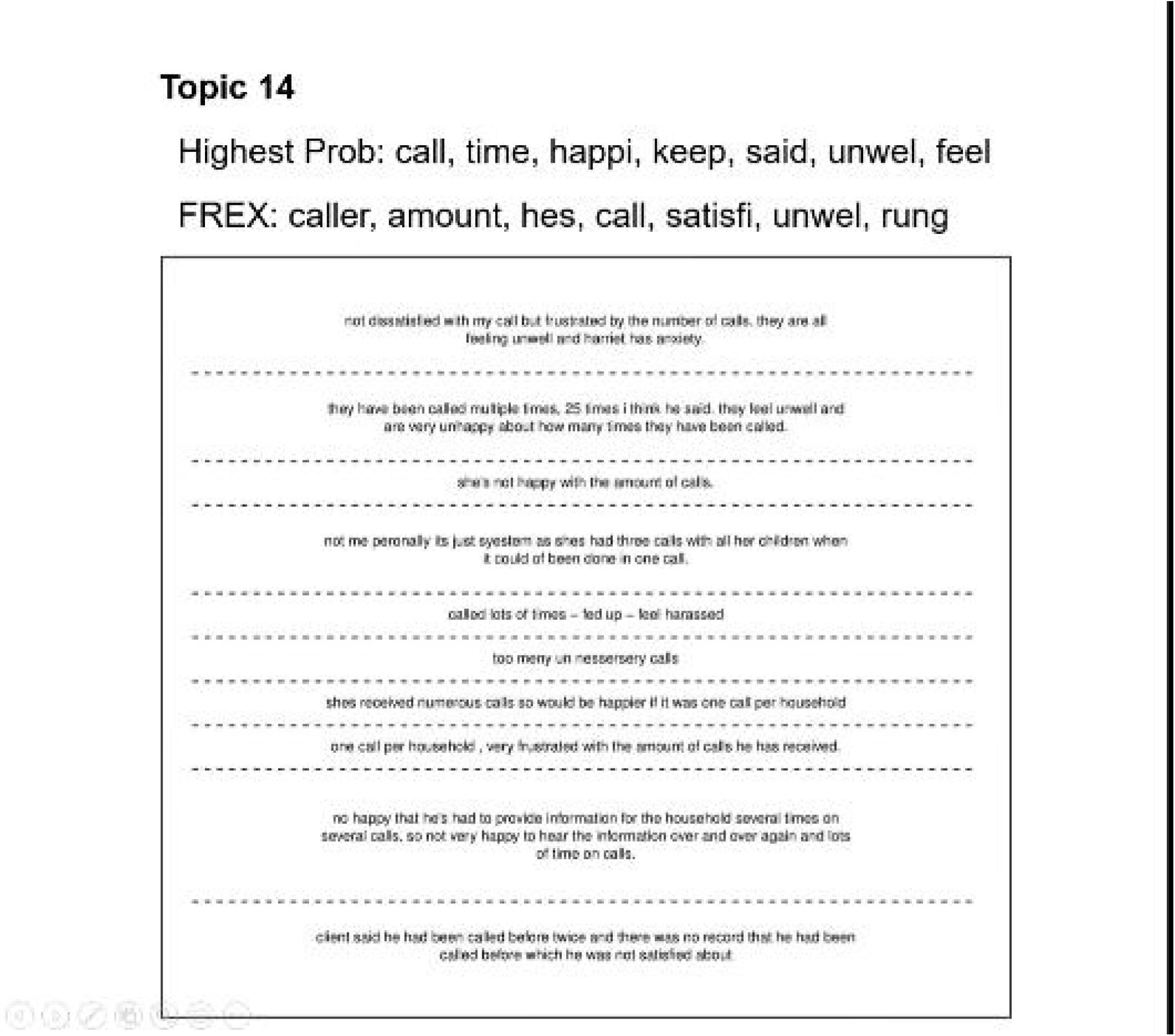
Diagram representing a framework for analysis of qualitative data for service improvement

### Strengths and limitations

First, the strength of this study relates to the use of the advancement in AI and applying the methods to analyse large volumes of unstructured data in public health emergency. Second, the systematically developed method which includes text analytics and qualitative data analysis demonstrated a proof of concept and paves the way for further application of this methods to various public health domains.

This study has several limitations. First, we have only analysed data provided by users in England and so our findings may not be generalisable to the rest of the UK or different contexts. In addition, these results might not be generalisable to those that disengaged with or who completed the Service but did not provide any feedback. This study does not provide an in-depth analysis and an exhaustive issue expressed by the users. However, the results are congruent with the qualitative analysis of the interviews conducted with the cases and contacts [21]. Furthermore, as we have limited the analysis to the feedback of users with dissatisfied responses, we cannot provide insight into the views of satisfied users around the same issues. A major issue with applying Structural Topic Modelling relates to the lack of control over the variables explored. Although this is one of the strengths of the method, it also limits the researchers’ ability to make strong inference about hypothesised results, making the method less robust to interpretation biases.

### Implications for research, practice, policy

#### Research

This study has several implications for research and further development of the model for automatic and pragmatic free-text data analysis. Specifically:

1. Further research is needed to develop a more sophisticated method that can be applied to monitor the changes in the Service over time.
2. As individuals have different experiences of the Service highlighting social inequalities, findings need to be stratified by sociodemographics
3. Need to develop a scalable model to apply it to accelerate understanding of different behaviours beyond the pandemic and into the post-pandemic recovery, namely the behaviours associated with poorer outcomes and higher mortality rates in those infected with SARS-Cov-2, such as unhealthy diet, physical inactivity, alcohol consumption and smoking.
4. The current model needs to be applied to different datasets in order to increase its generalisability and usability of the model for public health professionals.

#### Practice and Policy

There is a need to systematically reflect and gather the learnings from the pandemic in order to develop a more efficient response in the future. Some of the issues with the Service related to the usability and may be addressed in a time-efficient manner by simple technological changes. Some issues might require a more complex approach, for example beliefs about the inefficiency of the service. It is important that these issues are recognised and that various interventions addressing different problems are developed in order to developed a more efficient method of contact racing in the context of the pandemic.

#### Conclusion

This study provides a proof of concept of a method to analyse large quantities of free text data in service improvement. It includes an AI-enabled text analytics followed by a qualitative analysis conducted by a human. As such, this method has the potential to contribute to improvement of the Test & Trace Service, and other digital systems for public health safety or health improvement. More specifically, this method is best suited for cases where qualitative analysis is either needed rapidly or applied to large volumes of narrow in scope, short free-text responses. Importantly, this method is not suitable in cases where a nuanced qualitative analysis is the goal but can be a tool to augment and speed up existing qualitative approaches.

## Data Availability

All data produced in the present study are available upon reasonable request to the authors

## Declarations

### Ethics approval and consent to participate

This was a secondary analysis of data collected as part of the contact tracing process and so no formal ethics approval was sought.

### Consent for publication

Not applicable; no individual person’s data is included.

### Availability of data and materials

The dataset used in this research cannot be made available as it includes free-text data that potentially compromise individual privacy. There was no consent obtained for the data to be made publicly available. The code used to analyse the data has been made available in an online repository (doi:10.17605/OSF.IO/CVZD3).

### Competing interests

During the conduct of the study (data collection, analysis and interpretation), all authors were employed at Public Health England Behavioural Insights. From 1st October, PHE was dissolved, and all authors have been transferred to the newly created Office for Health Improvement and Disparities at the Department of Health and Social Care.

Neither PHE nor DHSC had any influence over the project or dissemination of the findings.

### Contributions

PB conceptualised the study, facilitated access to data and resources, identified and developed the methodology, conducted qualitative analysis and contributed to writing the original draft of the manuscript and editing later versions. TP developed the methodology, conducted quantitative and qualitative analysis of the data, contributed to writing the original draft of the manuscript and editing later versions. CS conceptualised the study, conducted preliminary data analysis and contributed to editing later versions of the manuscript. TC supervised all aspects of the study.

## Acknowledgements

We thank Charlotte Anderson and the team at Contact Tracing Cell: data and surveillance (Public Health England, now transitioned to UK Health Security Agency) for access to the data; we thank Julian Flowers (Public Health Data Science, Public Health England) for providing guidance on statistical methods; we thank Will Nicholson from Behavioural Science Team, Public Health England for helping us explore the various supervised methods for data analysis.

